# Seroprevalence of COVID-19 virus infection in Guilan province, Iran

**DOI:** 10.1101/2020.04.26.20079244

**Authors:** Maryam Shakiba, Seyed Saeed Hashemi Nazari, Fardin Mehrabian, Seyed Mahmoud Rezvani, Zahra Ghasempour, Abtin Heidarzadeh

**Affiliations:** School of health, Guilan University of medical sciences, Rasht, Iran; Cardiovascular diseases research center, Guilan University of medical sciences, Rasht, Iran; Department of epidemiology, School of public health and safety, Shahid Beheshti University of medical sciences, Tehran, Iran; School of medicine, Guilan University of medical sciences, Rasht, Iran; Health deputy, Guilan University of medical sciences, Rasht, Iran

**Keywords:** Asymptomatic Infections, COVID-19, Seroprevalence

## Abstract

**Background:** The extent of infection by coronavirus disease 2019 has not been well known. In this study we aimed to determine seropositivity of COVID-19 virus infection in population of a highly affected area in north of Iran.

**Methods:** In a population-based cluster random sampling design through phone call invitation, a total of 196 household including 551 subjects agreed to participate in this study. Each participant were taken 50ml blood sample at health care center. Rapid test kits were used to detect antibody against COVID-19. Crude, population-weight adjusted and test performance adjusted prevalence of antibody seropositivity to SARS-CoV-2 were reported.

**Results:** The prevalence of antibody seropositivity was 22% (95%CI: 19-26%). The population weight adjusted estimate was 21% (95%CI: 14-29%) and test performance adjusted prevalence was 33% (95%CI: 28-39%). Based on these estimates the range of infected people in this province would be between 518000 and 777000.

**Conclusion:** The population seropositivity prevalence of COVID-19 virus infection indicated that the asymptomatic infection is much higher than the number of confirmed cases of COVID-19. This estimate can be used to better detect infection fatality rate and decide for public policy guidelines.

## Introduction

Severe acute respiratory syndrome coronavirus 2 (SARS-CoV-2) that was first reported in China has now spread to all countries throughout the world. The disease that subsequently called as coronavirus disease 2019 (COVID-19) has an extensive spectrum of manifestation including asymptomatic infection, mild disease of upper respiratory system, severe viral pneumonia with acute respiratory syndrome, and death (1). Asymptomatic or subclinical infections are one of the major challenges of COVID-19 with public health concerns as it is presumed that they can spread the infection and remained undiscovered in the community (2-4). According to the report on “Diamond Princess”, 51% of all confirmed cases including 10 crew and 308 passengers were asymptomatic(5). The prevalence of asymptomatic infection in the community is unclear yet, however, this is important to be identified for at least two reasons. First, the unknown infection fatality rate that is an important public health measures of COVID-19, would properly be estimated by adequate knowledge of proportion of all persons infected with novel coronavirus (6, 7). Current fatality rate estimates are based on confirmed cases multiplied by a factor representing asymptomatic cases (8, 9). Moreover, for achieving the so-called temporary herd immunity, the minimum level of population immunity to halt the spread of infection in the community would be 1-(1/R_0_) (10). Where, R_0_ is the number of secondary cases generated by an infected individual in an ongoing epidemic and estimated to be 2 for Iran (11). Therefore, the information about the percentage of previously infected and hence immune people can help us to identify herd immunity, project epidemic and decide about public policy guidelines. According to World health organization (WHO), surveillance of antibody seropositivity including immunoglobulin G (IgG) and immunoglobulin M (IgM) against COVID-19 in a population can provide inferences about the extent and cumulative incidence of infection in the population (12). Guilan province was the second large city located at north of Iran that had several earliest number of COVID-19 soon after epidemic initiation in china. The epidemic curve in this province now has been stepped down making it an appropriate population-level location to test for the presence of active and past infections through seroprevalence survey. Therefore in this study we aimed to provide population-based seropositivity estimate of COVID-19 virus infection based on WHO protocol.

## Methods

### Study type and study population

This was a cross-sectional population-based study to investigate seroprevalence of COVID-19 virus infection in Guilan province, north of Iran in April 2020. The study had been approved by IRB of Guilan University of medical sciences, Rasht, Iran. Individuals permanently resided in Guilan province irrespective of age were eligible for recruitment into the study. All persons living in the household including children invited to participate in the study. Household were defined as at least two people living in the same place. Residence of Institutional living centers such as nursing homes, prisons, and boarding schools and those subjects who refused to give informed consent, were under active treatment of COVID-19, or had any contraindication to venipuncture were excluded from the study. With a prior prevalence of 50% for coronavirus infection with a 5% precision and design effect of 1.24, a total of 525 subjects were considered for sample size. Guilan province with a population of 2354848 is located in north of Iran and consisted of 16 counties. The study conducted in three high incidence counties (Rasht, Anzali, Lahijan) and two low incidence counties (Astara, Roudbar) according to the protocol of World health organization. A multistage cluster random sampling approach was used to select the participants. The number of clusters in each county was assigned proportional to the population size of that county. Therefore, 15 clusters were considered for Rasht which is the capital city of province, and 5 cluster for each of the remaining 4 counties. Clusters were identified at random. Then in each cluster, the household were selected using random digit dialing of mobile telephone number of head of the household that was previously registered in a community-based surveillance of coronavirus infection by health care centers.

### Data collection

A brief explanation of the research objectives followed by formal invitation of all family members to the study were given to the head of the household upon the phone call. For acknowledgment and to increase rate of participation a package of incentives including ethanol alcohol, face masks and kids incentives were considered for households. All invitation schedules were set to space enough to follow social distancing among households. At the day of attendance, after providing concise information about the risk and benefits accompanied by participation in this study, an informed consent was taken from the head of the household. An interviewer completed an electronic questionnaire containing participants’ demographic and exposure history information. Then, sample collectors in personal protective equipment drew 50 µL of capillary blood into an EDTA-coated microtainer. Tubes were labeled with participant ID. VivaDiag COVID-19 IgM/IgG from VivaChek was used for COVID-19-specific serological assay. According to manufacturer’s instruction, 10□µL of serum or whole blood sample was added into the sample port followed by adding 2 to 3 drops (70-100□µL) of dilution buffer. Test kits were read after about 15 minutes. According to the manufacturer, test performance characteristics showed a sensitivity of 97.1% in 11-14 days from infection and 81.25% in 4-10 days from infection. Test performance also verified on a sample of 30 PCR negative volunteer and 30 hospitalized patients. The sensitivity was 63.3%for both IgM and IgG (13). The specifity was reported to be 100% in both documents.

### Statistical analysis

Demographic characteristics described as frequency and percent. The crude age-specific population estimate of COVID-19 positive test were estimate as proportion of total sample size. Then, in order to account for 3-stage cluster sampling, survey design with probability weights equal to the inverse probability of selection of counties, clusters in each county and individuals in each cluster and also finite population correction for each stage was used. The cumulative incidence estimates were further adjusted for Rapid test sensitivity and specifity. We used Blaker’s methods to estimate standard error for cumulative incidence which account for the uncertainty in the sensitivity and specificity. Blaker’s method provide smaller exact two-sided confidence interval estimates (14). For test performance adjustment we applied the results of previously published sensitivity and specifity as 63% and 100%, repectively. The raw and the survey-weight adjusted estimates were performed in Stata version 13 (StataCorp LP, College Station, TX). Adjustment for test performance characteristics was performed using epiR package in R software.

## Results

From 632 household contact, 196 household including 551 subjects (49% male) were participated in this study. The major reasons for non-participation were concerning about getting infection in health facility center (17%), busy schedule (20%), lack of assurance to system (2%), no response (31%), and other non-specified reasons (28%). Table 1 provided demographic characteristic of unadjusted sample and population-weight adjusted of the sample and Guilan province estimates. The sample distribution was not meaningfully deviated from that of the province except for living place (32% in sample lived in village, 43% in the province).

**Table 1.** Demographic characteristics of sample relative to Guilan province estimates.

| characteristics | Unadjusted estimate<br>N(%) | Population adjusted<br>estimate | Province |
| --- | --- | --- | --- |
| Sex |  |  |  |
| Men | 270(49) | 0.49 | 0.5 |
| Women | 281(51) | 0.51 | 0.5 |
| Age group |  |  |  |
| < 5 | 27(0.05) | 0.04 | 0.06 |
| 5-18 | 107(0.19) | 0.19 | 0.17 |
| 18-60 | 343(0.62) | 0.60 | 0.63 |
| >60 | 74(0.13) | 0.16 | 0.14 |
| Living place |  |  |  |
| Village | 175(31.8) | 0.33 | 0.43 |
| city | 376(68.2) | 0.67 | 0.56 |
| Job |  |  |  |
| Employee | 57 (10) | .09 | Not reported |
| Housekeeper | 165(30) | 0.30 |  |
| Student | 122(22) | .21 |  |
| Unemployed | 68(12) | .12 |  |
| Farmer | 18(3) | .03 |  |
| Salesman | 47(8) | .09 |  |
| Health personnel | 44(8) | .07 |  |
| Driver | 13(2) | .02 |  |
| worker | 17(3) | .03 |  |

From total, 11 subjects could not be tested and 12 had invalid test results (i.e. test kit showed no control band). From remaining 528, 117 subjects were positive for either IgM or IgG, crude prevalence rate of 22% (95% confidence interval (CI): 19-26%). The population-weight adjusted prevalence estimates was 21% (95%CI: 14-29%). Table 2 presents the unadjusted, population weight adjusted prevalence estimates, and improved estimation by adjusting for test performance results. The adjusted test performance results yielded prevalence rate as 33% (95%CI: 28-39%).

**Table 2.**
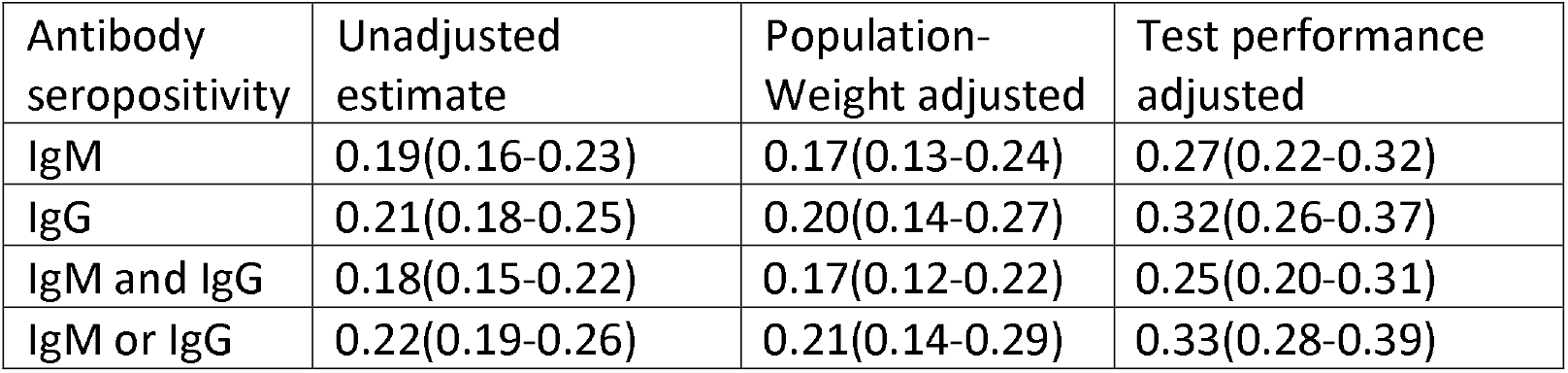
Seropositivity prevalence estimates in Guilan province.

**Table 3.** Seropositivity prevalence estimates according to the study variables.

| characteristics | Unadjusted | Population-weight adjusted |
| --- | --- | --- |
| Sex |  |  |
| Male | 0.22(0.17-0.27) | 0.19(0.13-0.26) |
| Female | 0.23(0.18-0.28) | 0.22(0.13-0.36) |
| Age group |  |  |
| < 5 | 0.15(0.06-0.35) | 0.16(0.04-0.46) |
| 5-18 | 0.20(0.13-0.29) | 0.16(0.08-0.31) |
| 18-60 | 0.23(0.19-0.28) | 0.21(0.14-0.30) |
| >60 | 0.26(0.17-0.38) | 0.26(0.15-0.41) |
| Place of residence |  |  |
| Village | 0.23(0.17-0.30) | 0.22(0.16-0.30) |
| town | 0.22(0.18-0.26) | 0.20(0.13-0.30) |
| Job |  |  |
| Employee | 0.36(0.24-0.49) | 0.34(0.16-0.59) |
| Housekeeper | 0.24(0.18-0.32) | 0.24(0.12-0.43) |
| Student | 0.20(0.14-0.28) | 0.18(0.11-0.29) |
| Unemployed | 0.16(0.09-0.27) | 0.15(0.07-0.29) |
| Farmer | 0.19(0.06-0.46) | 0.19(0.08-0.42) |
| Salesman | 0.11(0.04-0.24) | 0.10(0.03-0.26) |
| Health personnel | 0.29(0.18-0.45) | 0.26(0.11-0.50) |
| Driver | 0.38(0.16-0.67) | 0.29(0.06-0.73) |
| worker | 0.06(0.01-0.33) | 0.08(0.003-0.67) |
| County |  |  |
| Rasht | 0.25(0.20-0.31) | 0.24(0.19-0.30) |
| Anzali | 0.31(0.21-0.42) | 0.30(0.21-0.41) |
| Astara | 0.15(0.09-0.25) | 0.15(0.09-0.23) |
| Lahijan | 0.16(0.09-0.26) | 0.16(0.10-0.23) |
| Rudbar | 0.19(0.11-0.29) | 0.19(0.11-0.29) |

Age groups-specific prevalence rate showed that the highest prevalence of seropositivity was among subjects older than 60 years and children less than 5 years had the lowest seropositivity. There was no significant difference in seropositivity prevalence estimates in terms of sex, age group, place of residence, job and county of residential place. From total 528 participants who provided blood sample, 366 subjects (69%) had none of the symptoms attributed to COVID-19 including general, respiratory, or gastrointestinal symptoms. Among them, 18% (65 subjects) had positive antibody test results which indicate the percent of quite asymptomatic cases in our sample.

## Discussion

In this study, the seroprevalence estimate of SARS-COV-2 antibodies after adjusting for population and test performance characteristics was between 22 and 33%. The current results is much higher than previously seroprevalence estimate in California, USA which was between 2.49% and 4.16% (15). Our estimate is also higher than other preliminary findings in Italy (16) with 10%, Germany (17) with 14%, Colorado (18) point to 2% seropositivity, and New York city around 14% (19).

In this study test performance characteristic had the highest influence on reported estimates as test kit had lower sensitivity resulting in about 40% of false negative cases that might be missed by the test.

This study was among the first population-based seroprevalence study according to WHO protocol which incorporated cluster random sampling approach among community households of a highly affected area. In comparison to the estimates of western countries and even preliminary results of other provinces in our country, the high seroprevalence in our province may be explained by large amount of economic relationship with China in a free trade zone that is located at north of the Guilan province. The zone has been placed in Anzali that has the highest seropositivity prevalence in Guilan province.

Along with other seroprevalence surveys reported globally, the results of our study showed that the number of infection is much higher than the number of confirmed case reported based on PCR results. The community seropositivity prevalence may also represent the percentage of asymptomatic infection by novel coronavirus. In our study, the percentage of seropositivity in non PCR-confirmed or symptomatic COVID cases was 18% which may represent the rate of asymptomatic infection. The asymptomatic proportion of novel coronavirus infection in our study is also higher than previously reported SARS and MERS coronavirus epidemic which has reported between 0-7.5% (20-24).

Our seroprevalence estimate can be used for approximating infection fatality rate rather than case fatality rate. Based on the estimates of current study, the range of infected people in this province would be between 518000 and 777000. As of April 23 (the completion date of survey) 617 people have died of confirmed COVID-19 in Guilan province. This number corresponds to an infection fatality rate between 0.08-0.12% that is much lower than currently reported estimates of case fatality rate for COVID-19. The range of infection fatality rate in California was 0.12-0.2% (15).

Previous estimate of case fatality rate using lag time for fatality in china was between 0.25-3.0% (25) far higher than currently estimated infection fatality rate.

In our study, the household affirmative response rate was 31% and it may induce some sampling bias favoring those with prior COVID-like symptoms seeking antibody test. However, in our study only 11 subjects (2%) had a history of COVID confirmed diagnosis and 125 subjects (23%) had a combination of fever, dry cough, ache and tiredness as the major symptoms of COVID-19. Otherwise, bias favoring individuals in good health who are capable of participating in the test center may result in an underestimation of true prevalence.

## Conclusion

This study revealed that the prevalence of antibody seropositivity in a highly affected area was between 21 and 33%. The resulting number of infection in the population is much higher than the number of confirmed cases currently reported in Guilan province. These estimates have implications for future public policy guidelines and showed that the true infection fatality rate would be much lower than current estimates.

## Data Availability

Data are not publicly available.

## Acknowledgment

This research has been sponsored by Iran’s Ministry of health and performed under a research grant from deputy of research at Guilan University of medical sciences. The authors are grateful to all health centers’ employees and personnel for cooperating and conducting the survey as well as all participants that took part in this research.

